# Anxiety and depression among people living in quarantine centers during COVID-19 pandemic: A mixed method study from western Nepal

**DOI:** 10.1101/2020.11.05.20224436

**Authors:** BC Udaya Bahadur, Sunil Pokharel, Sabika Munikar, Chetan Nidhi Wagle, Pratik Adhikary, Brish Bahadur Shahi, Chandra Thapa, Ram Prasad Bhandari, Bipin Adhikari, Kanchan Thapa

## Abstract

**Background:** In response to the COVID-19 pandemic, incoming travelers were quarantined at specific centers in Nepal and major checkpoints in Nepal-India border. Nepal adopted a generic public health approaches to control and quarantine returnee migrants, with little attention towards the quality of quarantine facilities and its aftermath, such as the poor mental health of the returnee migrants. The main objective of this study was to explore the status of anxiety and depression, and factors affecting them among returnee migrants living in institutional quarantine centers of western Nepal.

**Methods:** A mixed method approach was used which included a quantitative survey and in-depth interviews (IDIs). Survey questionnaire utilized Beck Depression Inventory (BDI) and Beck Anxiety Inventory (BAI) tools, which were administered among 441 quarantined returnee migrants and IDIs were conducted among 12 participants which included a mix of quarantined migrants and healthcare workers from the quarantine centres. Descriptive and inferential analyses were conducted on quantitative data; and thematic analysis was utilized for qualitative data.

**Results:** Mild depression (9.1%; 40/441) and anxiety (16.1%, 71/441) was common among respondents followed by moderate depression and anxiety {depression (3.4%; 15/441), anxiety (4.1%, 18/441)} and severe depression and anxiety {depression (1.1%; 5/441), anxiety (0.7%, 3/441)}. Anxiety and depression were independent of their socio-demographic characteristics. Perceived fear of contracting COVID-19, severity and death were prominent among the respondents.

Respondents experienced stigma and discrimination in addition to being at the risk of disease and possible loss of employment and financial responsibilities. In addition, poor (quality and access to) health services, and poor living condition at the quarantine centres adversely affected respondents’ mental health.

**Conclusion:** Depression and anxiety among quarantined population warrants more research. Institutional quarantine centers of Karnali province of Nepal were in poor conditions which adversely impacted mental health of the respondents. Poor resources allocation for health, hygiene and living conditions can be counterproductive to the population quarantined.

## Introduction

Over the past few decades, there have been outbreaks of several zoonotic infections [1]. The recent pandemic of COVID-19 caused by a novel coronavirus emerged from China in December 2019. As of 5^th^ October 2020, there were 35,493,222 cases and 1,043,171 deaths reported, affecting 216 countries globally [2]. In the absence of effective pharmaceutical interventions for the prevention and control, non-pharmaceutical interventions such as physical isolation of cases and quarantine of contacts, in addition to the measures for social distancing, travel restrictions, wearing masks and improved hand hygiene, have been imposed globally in an attempt to curtail the spread of the disease [3].

As of 5^th^ October 2020, there were 89,263 positive cases and 554 deaths reported from Nepal [4]. In the Karnali province located in western Nepal, there were 3889 positive cases and 10 deaths reported due to COVID-19 [5]. As COVID-19 cases were rapidly increasing in the South Asian countries, institutional quarantine centers were established across Nepal to quarantine the travelers returning the country, especially across the southern border, amid the pandemic [6, 7]. In Karnali province alone, a total of 58,267 returning travelers were quarantined in various quarantine centers by the 5^th^ of October, 2020 [5].

With an increasing COVID-19 cases in India, Nepalese labor migrants started to return Nepal in high numbers [8]. A blanket public health approach to quarantine all the returning labor migrants before allowing them to go home was adopted to reduce the transmission. Institutional quarantine centers in Nepal were rapidly established as temporary quarantine facilities in pre-existing infrastructure such as schools, campuses, hostels, hotels, and other accommodating facilities [9]. However, their effectiveness was questionable from the beginning because of the compromise in quality over quantity of services. People were living in close proximity inside these quarantine rooms often sharing beds and other utensils, risking these facilities for outbreak hotspots [10]. Poor quarantine sites management leading to poor living conditions in relation to basic needs, water, hygiene and safety were prominently identified [6, 11, 12].

Previous reports have found negative psychological impacts among quarantined people with longer duration of quarantine, fear of infection, frustration and boredom, inadequate supplies, and inadequate information as common stressors [13]. The trade-off between compromise in psychosocial health and complacency that these quarantine centres (as a symbol of public health measures against COVID-19) offer is an important paradox that needs urgent attention. The impacts of these control measure can have equal if not more adversities than the COVID-19 itself [14].

Although few studies in the past have addressed the psychological impact of quarantine (facilities) among population, they were mostly conducted in the high income settings, focused on health professional and non-migrant residents, and explored the impact of quarantine in pre-COVID-19 scenarios [13, 15–17]. Little is known about mental health outcomes among vulnerable population such as returnee migrants who were obliged to stay in quarantine centres before going home. This study aims to explore the status of anxiety, depression in the returnee migrants in institutional quarantine centers of western Nepal and factors that influence their mental health conditions.

## Methods

### Study Design

This study utilized a mix of quantitative and qualitative approaches among returnee migrants and health workers in quarantine institutions of Karnali Province in western Nepal to explore the status and drivers of anxiety and depression. The study followed a standard cross-sectional design using STrengthening the Reporting of OBservational studies in Epidemiology (STROBE) Checklist (Appendix 1) for quantitative part and COnsolidated Criteria for REporting Qualitative research (COREQ) guideline for qualitative aspect of the study (Appendix 2).

### Study population and study area

Nepal is divided into seven provinces where the administrative structure consists of federal, provincial, district and local level facilities [18]. This study was conducted in nine selected quarantine centers of Surkhet district, the capital of Karnali province in the western Nepal (Fig 1). After the outbreak of COVID-19 worldwide, the quarantine centers in Nepal were set up and managed by the provincial and local governments. People entering to Karnali province were quarantined for a minimum of 14 days upon their arrival before they could go home. Most of the people in these quarantine centers were labor migrants returning from India [19], who crossed the border by road as the cases of COVID-19 was rising and India imposed nationwide lockdown [7]. Health workers were mobilized to provide health-related support in these quarantine centers.

**Fig 1.**
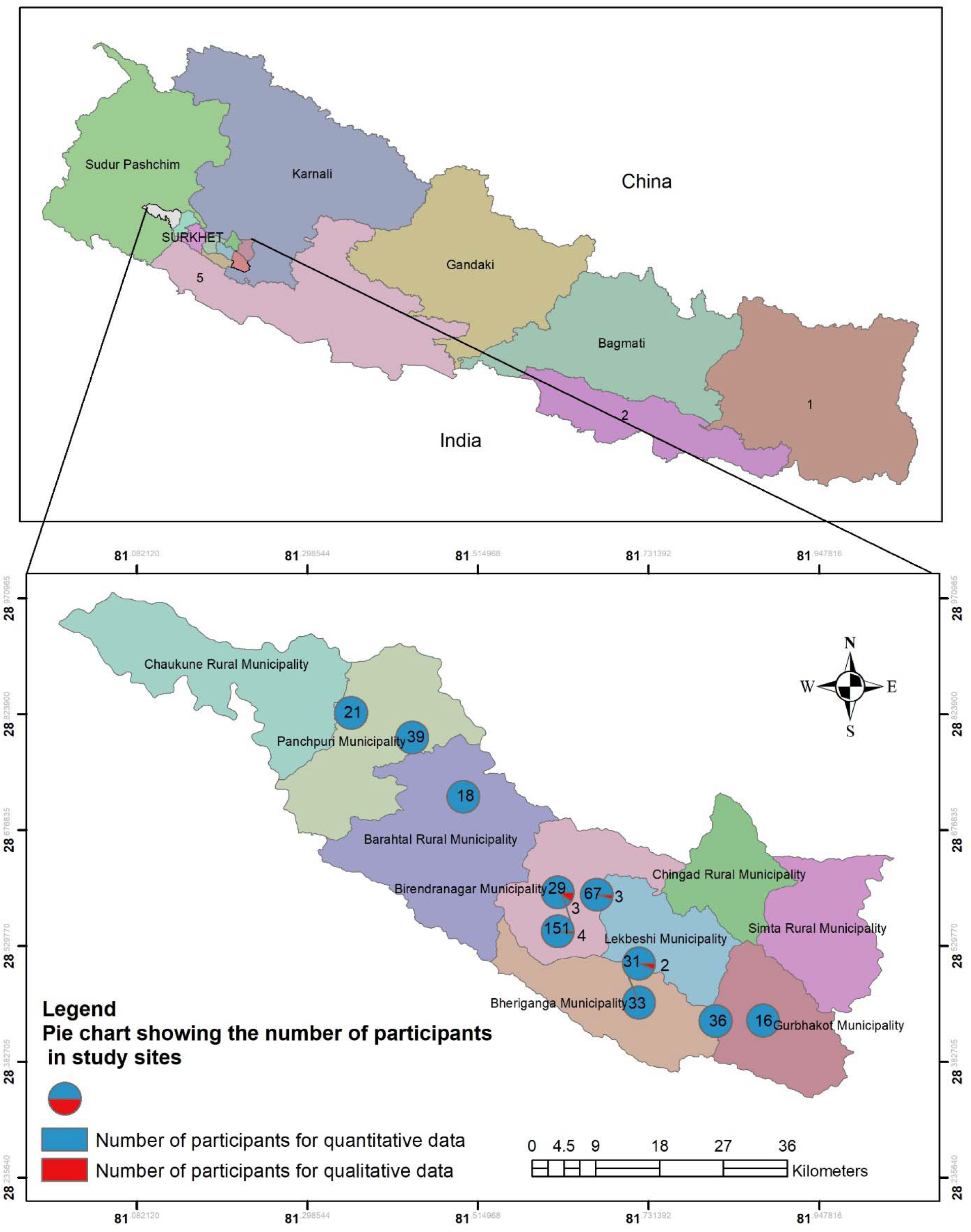
Study sites with number of participants in Surkhet district, Karnali province, Nepal. The map was created with ArcMap version 10.8.1 and GPS coordinates were taken from field locations.

Returnee migrants residing in various quarantine centers of Surkhet were approached for participation in the study. The data collection team purposively recruited the participants for quantitative information and IDI. A verbal informed consent was obtained from each participant before enrollment into the study.

The population sample for data collection was chosen from the available quarantined people who were staying in quarantine centres between 21^st^ April and 15^th^ May 2020. A total of 441 people participated in the questionnaire survey. IDI was conducted among 12 participants, which included six quarantined migrants and six health workers. Number of respondents (sample size) for IDI was based on the principles of ‘data saturation’, that is data were collected until no new data/themes emerged from further interviews [20].

### Data collection

Health workers working in the quarantine sites were trained for data collection using survey questionnaire. Face-to-face interviews were conducted with the quarantined people that lasted approximately 20 to 30 minutes. Survey questionnaire was used to obtain socio-demographic information of participants, and their status of anxiety and depression using Beck Depression Inventory (BDI) and Beck Anxiety Inventory (BAI) tools [22, 23]. The BDI and BAI questionnaires are validated and reliable tools in the context of Nepal [24, 25]. Both the BDI and BAI tools consist of 21 items with scale ranging from 0 to 3 in likert scale for each item and total score for both the tools ranges from 0-63. We used the scores of 0-13 to denote minimal depression, 14-19 for mild, 20-28 for moderate and 29-63 for severe depression in BDI scale, as implemented previously [26]. Similarly, the scores of 0-7 reflected a minimal anxiety, 8-15 reflected mild anxiety, 16-25 reflected moderate anxiety and 26-63 reflected severe anxiety [27].

The IDIs allowed to explore the issues and topics identified through quantitative questionnaire. Face-to-face IDIs were conducted among quarantined people and health workers by UBB (MA) and KT (MPH, MPhil) using thematic guide. Both UBB and KT are male public health experts in Nepal with over 10 years of experience in qualitative social science research. A brief informal discussion was conducted with each participant before interviews to establish relationship and facilitate participation. Participants were informed about the aims and objectives of the study. All the participants approached for study agreed to participate and there was no drop-out. Each of the IDIs lasted for approximately an hour and was audio-recorded. No repeat interviews were conducted with any participants. Field notes were made by the interviewers during the study.

All the quantitative and qualitative interviews were conducted directly with participants at convenient locations in the quarantine centers, talking necessary precautions against COVID-19 transmission. The survey questionnaire and IDI guide were designed in English language and translated into Nepali language. The tool was translated back to English for validation before administration. The survey questionnaire was pretested among 10% of the total sample size which were excluded from final analysis. Similarly, the IDI guide was pretested among 2 participants prior to its administration for final data collection.

### Ethical consideration

This study obtained ethical approval from Nepal Health Research Council dated 20^th^ April 2020 (Reference #2194). The interviews were conducted in Nepali language. A verbal informed consent was obtained from each participant and the consent was audio-recorded for IDIs. Approval from their immediate care takers were taken in case of those with age < 18 years for recruitment in this study. No financial incentive was given to the participants for their participation.

### Data analysis

The quantitative data were entered in MS Excel 2019 and then transferred into SPSS Version 23 for analysis. Descriptive and inferential analysis were made against the scores for depression (BDI) and anxiety (BAI). Chi-square statistic was used to test for the association of depression and anxiety with socio-demographic characteristics of the population. For qualitative data, content analysis was conducted to derive and synthesize themes. The IDIs were transcribed into Nepali and translated into English. The transcripts were collated in MS word. Thorough desk review and coding of the data was done by two investigators SP and KT adhering to the research question. Data were first categorized into themes using a deductive approach based on the thematic guide and were developed based on the initial coding of the transcripts. Emerging themes were added after reading each new transcript using inductive approach. Excerpts were chosen based on their relevance to the research questions, and thematic prominence. No participant feedback was sought on the findings. Triangulation of data from quantitative and qualitative methods and among the two types of respondents (quarantined migrants and health workers) was done for the analysis.

## Results

### Study participants and background characteristics

Among 441 participants, 282 (63.9%) were aged 18-36 years and majority (90%; 395/441) of the respondents were labor workers. Only a minority (2%; 8/441) of the participants had higher education (>12 grades) and majority (93%; 410/441) of the participants earned less than 200 USD per month (Table 1).

**Table 1.** Socio-demographic characteristics of participants in quantitative survey (n= 441).

| Characteristics | Frequency(n) | Percent (%) |
| --- | --- | --- |
| **Age (Years)** |  |  |
| <18 | 58 | 13.2 |
| 18-35 | 282 | 63.9 |
| 36-53 | 88 | 20.0 |
| >54 | 13 | 2.9 |
| **Educational status** |  |  |
| Ill-literate | 39 | 8.8 |
| Incomplete Basic | 123 | 27.9 |
| Secondary | 271 | 61.5 |
| Higher Education (>12 Class) | 8 | 1.8 |
| **Employment** |  |  |
| Labor | 395 | 89.6 |
| Self Employed | 27 | 6.1 |
| Hotel Staffs | 7 | 1.6 |
| Service | 5 | 1.1 |
| Driver | 4 | 0.9 |
| Security Guard | 2 | 0.5 |
| Musician | 1 | 0.2 |
| **Monthly income** |  |  |
| Less than 10 thousand NPR | 21 | 4.8 |
| 10-20 Thousand NPR | 389 | 88.2 |
| More than 20 thousand NPR | 31 | 7.0 |
| **Household size** |  |  |
| <4 | 132 | 29.9 |
| 5 – 9 | 279 | 63.3 |
| >10 | 30 | 6.8 |
| **Use Social media** |  |  |
| Yes | 256 | 58.0 |
| No | 185 | 42.0 |
| **Have a Bank Account** |  |  |
| Yes | 109 | 24.7 |
| No | 332 | 75.3 |

Mean Income (NRs):12380.92, Average Household Size =5.76

### Status of depression and anxiety

Around 13.6% (60/441) of individuals kept in quarantine centres were suffering from depression (Mild: 9.1%; 40/441, Moderate: 3.4%; 15/441, and Severe: 1.1%; 5/441). Slightly higher proportion (20.9%, 92/441) of respondents were suffering from anxiety (Mild: 16.1%; 71/441, Moderate: 4.1%; 18/441) and Severe: 0.7%; 3/441) compared to depression (Table 2). There was no significant association of depression and anxiety with socio-demographic characteristics of the population (Table 3 and Table 4).

**Table 2.** Status of depression and anxiety level (n= 441).

| **Depression level** | **Frequency (n)** | **Percent (%)** |
| --- | --- | --- |
| Minimal | 381 | 86.4 |
| Mild | 40 | 9.1 |
| Moderate | 15 | 3.4 |
| Severe | 5 | 1.1 |
| **Anxiety level** |  |  |
| Minimal | 349 | 79.1 |
| Mild | 71 | 16.1 |
| Moderate | 18 | 4.1 |
| Severe | 3 | 0.7 |

**Table 3:** Relationship between depression and independent variables.

| **Variables** | **Category** | **Depression** |  | **Chi-square value** | **p- value** |
| --- | --- | --- | --- | --- | --- |
|  |  | **Minimal** | **Depression** |  |  |
| **Age** | <30 years | 238 (85.3%) | 41 (14.7%) | 0.76 | 0.472 |
|  | ➤ 30 years | 143 (88.3%) | 19 (11.7%) |  |  |
| **Education** | Illiterate | 29 (74.4%) | 10 (25.6%) | 5.79 | 0.055 |
|  | Basic | 110 (89.4%) | 13 (10.6%) |  |  |
|  | Secondary and above | 242 (86.7%) | 37 (13.3%) |  |  |
| **Income** | <15000 | 322 (87.0%) | 48 (13%) | 7.82 | 0.351 |
|  | >15000 | 59 (83.1%) | 12 (169%) |  |  |
| **Household size** | <4 | 213 (86.6%) | 13 (13.4%) | 0.017 | 0.890 |
|  | >4 | 168 (86.2%) | 27 (13.8%) |  |  |
| **Occupation** | Labor | 343 (86.8%) | 52 (13.2%) | 0.626 | 0.493 |
|  | Other | 38 (82.6%) | 8 (17.4%) |  |  |
| **Have social account** | Yes | 220 (85.9%) | 36 (14.1%) | 0.108 | 0.780 |
|  | No | 161 (87.0%) | 24 (13.0%) |  |  |
| **Have bank account** | Yes | 91 ( 83.5%) | 18 ( 16.5%) | 1.042 | 0.334 |
|  | No | 290 (87.3%) | 42 (12.7%) |  |  |

**Table 4.** Relationship between anxiety and independent variables.

| **Variables** | **Category** | **Anxiety** |  | **Chi-square value** | **p- value** |
| --- | --- | --- | --- | --- | --- |
|  |  | **Minimal** | **Anxiety** |  |  |
| Age | <30 years | 228 ( 81.7%) | 51 (18.35) | 3.06 | 0.089 |
|  | > 30 years | 121 (74.7%) | 41 (25.3%) |  |  |
| Education | Illiterate | 29 (74.4%) | 10 (25.6%0 | 0.851 | 0.0653 |
|  | Basic | 96 (78.0%) | 27 (22.0%) |  |  |
|  | Secondary and above | 224 (80.3%) | 55 (19.7%) |  |  |
| Income | <15000 | 289 (78.1%) | 81 (21..9%) | 1.477 | 2.66 |
|  | >15000 | 60 (84.5%) | 11 (15.5%0 |  |  |
| Household size | <4 | 193 (78.5%) | 53 (21.5%) | 0.157 | 0.724 |
|  | >4 | 156 (80.0%) | 39 (20.0%) |  |  |
| Occupation | Labor | 38 (78.0%) | 87 (22.0%) | 3.106 | 0.86 |
|  | Other | 41 (89.1%) | 5 (10.9%) |  |  |
| Have social account | Yes | 202 (78.9%) | 54 (21.1%) | 0.020 | 0.906 |
|  | No | 147 (79.5%) | 38 (20.5%) |  |  |
| Have bank account | Yes | 85 (78.0%) | 24 (22.0%) | 0.117 | 0.786 |
|  | No | 264 (79.5%) | 68 (20.5%) |  |  |

### Characteristics of study participants in IDIs

A total of 12 respondents, six each quarantined migrants and health workers from various quarantine centres participated in the IDI. Age of participants ranged from 22 years to 46 years. Immigrants were quarantined in different centres both in India and Nepal, the total duration ranging from 9 to 45 days. Health workers with various educational backgrounds were recruited in this study.

### Factors influencing mental health

#### Overall findings

Mental health of people living in quarantine centers were influenced by multiple factors (Fig 2). People living in quarantine mostly feared about disease and death, thought about health of themselves and family members, and their financial responsibilities. Discrimination and stigmatization by the community for being migrant and at higher risk of COVID-19 and poor quality of quarantine centers worsened their physical as well as mental health.

**Fig 2.**
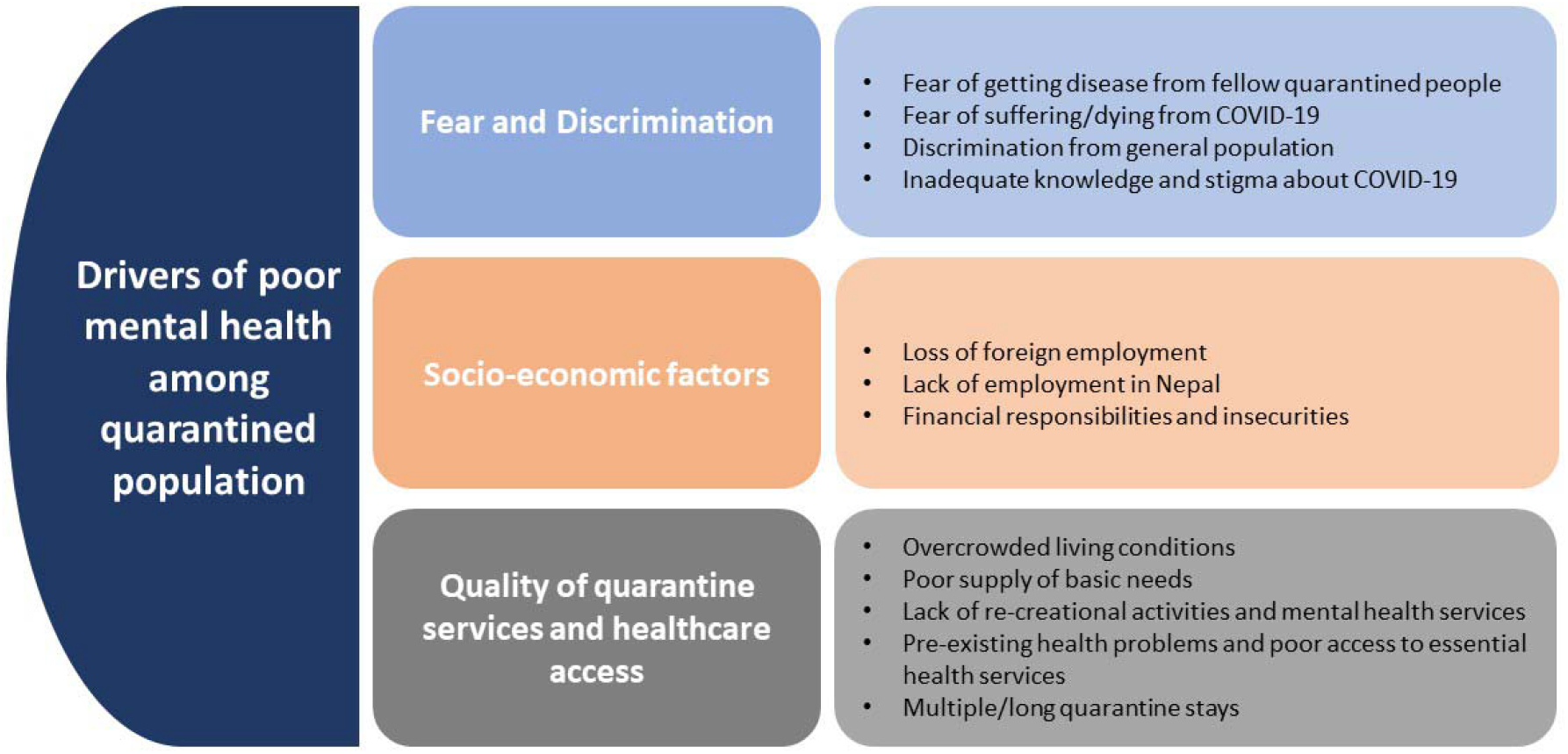
Summary of factors driving poor mental health in quarentine centers

#### Fear and discrimination

Almost all the participants were aware of the mode of disease transmission, common clinical presentation of the disease, and the role of social distancing, the use of masks and sanitization in reducing transmission. They also expressed their strong commitment to stay in quarantine to save their family and the community from acquiring disease from them.

> *“…This disease is highly contagious. Even my friends who travelled along with me are infected however it can be prevented using masks and frequent handwashing. This is an emergency condition and we [family and community] are at risk. So, we need to be safe*.*”*
>
> [IDI-6]

Almost all participants were also aware of the rising case and death tolls due to COVID-19 globally. The disease was perceived to be extremely contagious and the fear of dying from the disease was widely prevalent. Although, all of them were asymptomatic for COVID-19, they expressed their worry of contracting the disease from the fellow quarantined people they had to live in close proximity with. Some of the respondents were also aware of their fellow travelers testing positive for the disease and the high rate of disease transmission in Indian cities where they worked before travelling to Nepal which escalated their fear of being infected although were currently symptomless.

> “*One of our friends returning to Nepal from India has been diagnosed positive in Dailekh So I feel worried whether my reports also come positive*.”
>
> [IDI-3]

Respondents also expressed their dissatisfaction towards the authorities mixing them with more in-coming people which they thought could increase their risk of infection. This was often the reason for conflict between quarantined people and the authorities, and newcomers and people residing earlier in quarantine.

> “….*we did protest to not let 25 newcomers stay here. The local people also protested and threw some stone towards them at midnight [but it didn’t work]*.”
>
> [IDI-5]

The quarantine centers in Nepal were rapidly set up in response to a large number of migrants coming from India and operated without adequate measures to prevent transmission within the centers. Travelers were provided space based on the availability and people coming from different places and at different times were often sheltered in the same rooms.

The fear of contracting the disease, suffering and dying from the disease stirred the mental well-being of the participants which was often expressed in the form of restlessness and insomnia.

> *I sleep only at around 2-3 am and get up early in the morning. Sleep is disturbed and I am worried if I will go ill. I also get irritated when someone speak to me while I am busy doing other things*.
>
> [IDI-3]

Some participants associated their fear to the hyped-up uncensored news in the media and rumors about the disease which also enabled them to ruminate about the disease and potential suffering. Participants could see the impacts of hyped-up news and misinformation related to immigrants that may have led to high discrimination towards them. Despite that most were asymptomatic, they still faced the acts of discrimination. The discrimination was based on, in addition to the fear of them bringing the disease as they were returning from high transmission areas in India, the political and socio-economic factors. They were often denied food and water during their journey.

> *“The people from province 5 did not even give us water [for being from another province]*.*”*
>
> [IDI-6]

Participants were vocal about the discrimination faced by them and often expressed their annoyance towards these acts that hurt their esteem. They compared their quarantine experience in Nepal with their travel in India while returning home and expressed their dissatisfaction with the management of quarantine centers and the behavior of authorities towards them.

> “*How can you just undervalue us and provide just a packet of noodles and some water for a meal?*
>
> [IDI-12]
>
> *“Whenever we ask for something, the thing is thrown towards us rather than passing it on hands. In India, we used to stay in queue and get required articles respectfully*.
>
> [IDI-5]

#### Socio-economic factors

Participants were mostly labor migrants to India, most of them with a monthly income of less than $200. They had lost their job due to the pandemic and were returning home without a saving. Most of them reflected on their expectations of going to India and how it was troublesome for them to return without an earning.

> “*In my family, I was supposed to earn some money. I went to India thinking so. Now, I am returning without a penny in the pocket*.”
>
> [IDI-3]

Some participants further related their worsening economic condition with their family responsibility and expressed their concern over earning for living and providing education to their children in Nepal.

> “*I have a young child and another child is about to go to school. Now I don’t even have a single cent of money. Because of this, I think about my family. What they have been doing? I have been staying for such a long period. What children might have eaten at home? I am now unable to afford my children’s education*.*”*
>
> [IDI-3]

India is a popular destination for the labour migrants, especially from hilly regions in western Nepal. Among all the labor migrants, most are seasonal travelers who work for few months in Indian labor market each year to compensate the insufficient agriculture-based economy while some stay for longer duration to support their families in Nepal. Participants expressed that their land in the village were infertile to alone sustain their living for the whole year and voiced their insecurities for finding an employment in Nepal.

#### Quality of quarantine services and healthcare access

Almost all the participants reported an unpleasant quarantine experience in terms of their living condition, foods provided, care and treatment they received from the quarantine sites. The poor living condition at the quarantine centers were reported to be one of the stressors by many participants.

> *“…packaged meals used to be delivered in quarantine centre for us, but the food used to be left unattended in the open ground. We felt being treated like dogs*.*”*
>
> [IDI-5]

Poor facilities for water and sanitation deprived them from the maintenance of their personal and environmental hygiene. In addition to these, being a factor for their personal discomfort and dissatisfaction, this was disturbing the residents living around the quarantine centres and was often a reason for complains against quarantined population.

> “…*an auntie here [community person housed next to quarantine centre] complains that we are being unhygienic and dirty. But we have problem with water here, we are not given enough water. We haven’t been able to take bath for several days*.”
>
> [IDI-3]

People were often quarantined at more than one place in Nepal and India during their travel. For many participants, this led to very long cumulative quarantine duration, which was reported to be up to 45 days by a participant. Their physical and mental suffering while spending time in these various centres, with different group of fellow travelers and under management of various provincial and local authorities were reported to be very painful and annoying. A participant expressed his frustration over the physical torture from the authorities during the travel and at the quarantine sites.

> “*While staying in [quarantine sites] India, I was beaten by the police for requesting them to send me to Nepal. Later, after spending 45 days [quarantined in various centers in India], we were taken to Nepal at the mid-night*.?”
>
> [IDI-3]

There were no rooms for any re-creational activities. The fellow quarantined people were just the ‘strangers’ for most of the participants. When asked if they got chance to share their problem and experiences with other people in the quarantine, they expressed their difficulty to open up with anyone who were strangers to them. Although quarantined people were aware of the need to quarantine them; and complied with the government regulation to stay in the quarantine center for at least 2 weeks after arrival in Nepal, they thought they were living a miserable life there.

> “*Thinking about the family and home, I wish I could escape and run away from here now*.*”*
>
> [IDI-6]

Although some quarantine centers had started some forms of mental health services such as yoga, the participants explained that counselling services in quarantine centers were poor (worse than the expected).

> *“All quarantined people are worried and stressed. No one is there to counsel. We should cope with ourselves and decide what should be done to make the situation better*.*”*
>
> [IDI-3]

Participants also expressed lack of confidence in the health system of the country when COVID-19 was spreading. Many of them compared with the health services in India where they went through rapid test for COVID-19 screening and expressed their displeasure as the quarantine centres in Nepal merely tested for their body temperature. They thought there were no facilities for optimal routine health services at the quarantine centres and were worried of encountering health crisis during their stay.

Many participants had chronic problems such as gastritis, diabetes and hypertension and they felt the need to follow-up with health professionals for their medications. Some participants felt that their pre-existing health conditions were worsening due to the lack of proper consultation and treatment. Although some health workers thought that they were providing the essential services they needed, other accepted that the available health services in the quarantine centres were limited.

> *“One day, an aunty [a quarantined female] came crying to our tent [health support unit] and told me about her problems. She was also a diabetic patient and had developed rashes over her body. She showed her rashes and was also worried about her sugar control. Then, we gave her medicine for rashes, but could not test for her blood sugar. We had to counsel her to stay in quarantine and defer her sugar testing until she completed her quarantine duration*.*”*
>
> [IDI-12]

## Discussion

The main objective of this study was to explore the status of mental health and factors affecting it among returning labor migrants staying in institutional quarantine centres in western Nepal during COVID-19 pandemic. While blanket public health approach in Nepal was directed towards the isolation and quarantine of people coming across the border for the prevention of COVID-19 transmission, much less attention has been paid towards the mental health status of the people who stayed in the institutional quarantine centres.

### Prevalence of anxiety and depression

This study reported prevalence of anxiety to be 20.9% and depression to be 13.6% which aligns with a previous reports that highlights high prevalence of depression and anxiety in population affected by COVID-19 ranging from 16-28% [28]. A significantly higher prevalence of anxiety and depression among those affected by quarantine (12.9% and 22.4%) compared to those unaffected (6.7% and 11.9%) has been reported from China [29]. Some studies have reported alarmingly high occurrence of mental disorders among those being quarantined. For instance, among Filipino quarantined respondents, 60.3% had moderate to severe anxiety and 53.1% had moderate to severe depression [30]. Although large heterogeneity in the tools used in the evaluation of mental illness exists across various studies and contextual factors are in effect, isolation and quarantine increases the vulnerability of population to mental health problems needing a serious attention.

While emerging evidence indicate increased anxiety and psychological distress due to COVID-19, factors other than disease itself significantly affects the mental wellbeing of the population. For example, in a study from Germany where over 50% of participants expressed anxiety and psychological distress associated with COVID-19 pandemic; psychological and social determinants showed stronger associations with anxiety and distress rather than experiences with the disease [31].

### Factors affecting mental health

Perceived COVID-19 threat itself is a recognized risk factor for depression and anxiety; and has been reported to trigger metal health problems in general population [32]. Increasing number of reported deaths worldwide triggers increased psychological distress among population further progressing to mood disorders [33]. The fear of suffering and dying from COVID-19 was highly prevalent among the quarantined population. The fear was further aggravated by the news of increasing cases and death tolls. The uncensored and often exaggerated news in media were other enabling factors for the participants to ruminate about the disease and potential suffering and have prompted fear and discrimination among general population [34]. Participants reported discrimination either from peers, health workers and/or community people. Perceived discrimination among quarantined people is conducive to higher mental health distress [35].

Economic challenges are emerging worldwide manifesting in the forms of loss of job, increased responsibilities and financial loss leading to increased stress level [8]. India is a popular destination for the labor migrants, especially from hilly regions in western Nepal [36–38]. These labor migrants are either seasonal travelers who work for few months in Indian labor market each year to compensate the insufficient agriculture-based economy or employee for a longer duration to support their families in Nepal. Loss of jobs in Indian labor market and decreasing job opportunities in Nepal [39–41] with the evolving pandemic relates to the impending economic crisis in the lives of population with minimal education background and very low economic status as reported in this study. World Bank emphasizes on a significant impact of COVID-19 on labor market [42] which has increased risk of depression associated with greater financial difficulties [43]. In china, higher levels of anxiety and depression is reported among people with severe economic loss [29].

Poor access to health care services is reported from the quarantine centers in Nepal [6, 10]. As the health system in Nepal was already strained due to the impact of ‘covidization’, people lacked access to essential health services [34, 44]. Many of the participants already had medical conditions which needed urgent care. People with self-perceived illnesses supposedly have higher depression and anxiety level in the absence of proper consultation and care. The insufficient health infrastructure and health manpower in the country to handle the COVID-19 cases and other essential health services were questioned, but no adequate efforts were made to strengthen the health system capacity [21]. One-third of the available inpatient beds and ventilators were allocated for the treatment of covid-19 cases [9] however, with existing limited functional capacity in the country, this could jeopardize both COVID-19 management and essential health services.

Poor infrastructure of quarantine centers and lack of proper supplies including food, water sanitation and hygiene contributed for poor living condition of people in the quarantine centers [6, 10]. These factors not only increases risk of COVID-19 outbreaks within quarantine centers [10], and acquiring various other infections [45], they largely contribute to the poor mental wellbeing of the people. Some participants reported increased duration of quarantine stay making them more vulnerable to stress and mental problems. Adequate psychological supports and mental health services to quarantined people are required to deal with their mental health difficulties [46]. The provision of recreational facilities, counseling services and peer support were recognized as the key factors to improve the living conditions of people in the quarantine centers and enhance their mental wellbeing[47, 48].

Imposed strict quarantine may have debilitating effects in mental health. Psychological impact of quarantine is pervasive, substantial and can be long lasting [13]. In parallel to instituting essential quarantine measure to curb the spread of COVID-19, it is urgent to think about psychological needs of those people being quarantined and health workers responding to the situation [21, 49]. The psychological consequences during disasters are often the neglected aspects in public health preparedness [50] and was reflected in Nepal’s current COVID-19 preparedness too [51]. The psychological aspects in a disaster should be of paramount importance with preparedness in place to mitigate the long term aftermath: anxiety and depression [52].

### Strengths and limitations

This is to our knowledge the first study conducted in Nepal to explore anxiety and depression among people who had to stay in the quarantine centres during COVID-19 pandemic. One of the main strengths of this study is that it uses a mixed method design in an adequate sample. Nonetheless, the study may have incurred recall and social desirability bias. In addition, anxiety and depression in this study may have accumulated from the contribution of various previous circumstances apart from quarantine experience. Other quarantine centres in Nepal may have different characteristics in terms of services and ambience possibly impacting the level of mental wellbeing. However, poor services, and crowded living conditions have been reported through from across Nepal through media. Future research can explore into the nationwide situation of quarantine centres and their impacts on the mental wellbeing population quarantined in these centers.

## Conclusion

High prevalence of anxiety and depression was reported among returnee labor migrants living in institutional quarantine centers of Karnali province of Nepal amid COVID-19 pandemic. Although no statistically significant association of anxiety and depression with socio-demographic variables was observed, the qualitative approach identified various factors to contribute for their poor mental health. Institutional quarantine centers were in poor conditions in relation to basic supplies, health, hygiene and recreational support which adversely impacted mental health of the respondents. Measures towards alleviating fear and stigma and ensuring financial securities of the population during the time of health crisis are important for preparedness against epidemics such as COVID-19.

## Recommendations

- Programmatic response to COVID-19 with efforts towards the containment of disease using measures for the quarantine of high-risk population should in parallel prioritize the mental wellbeing of the quarantined population.
- Steps towards alleviating fear and stigma through education and awareness about the disease should be implemented.
- Censoring exaggerated and unverified news and ensuring that the information being disseminated through media is factual and reliable is necessary to mitigate the damage made by fear on the mental wellbeing of the population.
- Strategies for promoting societal acceptance and psychosocial support are very crucial to mitigate the impact of stigma and discrimination.
- Healthcare preparedness should incorporate mental health services including proper psychological counselling services and adequate essential health services for the vulnerable quarantined population.
- Attention needs to be paid to the quality of life in quarantine centers with sufficient basic and recreational support.
- Healthcare budgeting should address the financial needs of population with low economic status at the time of healthcare crisis and make pre-emptive preparation for providing financial support and ensuring financial security of the vulnerable population in need.

## Data Availability

All quantitative data pertaining to this study are presented in the study. Full details of qualitative data presented in this study are restricted for sharing because of the nature of breaching the anonymity due to quotes and expressions which are identifiable.

## Abbreviations

ANM: Auxiliary Nurse Midwife
BAI: Beck Anxiety Inventory
BBS: Bachelor in Business Studies
BDI: Beck Depression Inventory
BPH: Bachelor in Public Health
HA: Health assistant
HQ: Home quarantine
HW: Health worker
ML: Migrant labour
MoSD: Ministry of Social Development
SB: Street Business

## Declarations

### Authors’ contributions

UBB and KT conceived the idea and designed the study; UBB, CNW, CKT, RPB and KT conducted field studies and acquired the data; SP, SM, BA and KT analyzed the data; SP, SM and KT wrote the first draft; all the authors critically revised the manuscript and agreed on the final version of the manuscript.

### Consent for publication

Not applicable

## Acknowledgements

We would like to thank all the colleagues at the Public Health Service Office, Surkhet for support throughout the study. We would also like to thank Mr Dev Ram Sunuwar from Armed Police Force of Nepal for creating the map (Fig 1) for the study.

